# Impact of the Food4Moms Produce Prescription Program on Readiness for Healthy Eating, Fruit and Vegetable Intake, and Food Security

**DOI:** 10.64898/2026.04.24.26351720

**Authors:** Sofia Segura-Pérez, Katina Gionteris, Amber Hromi-Fiedler, Kathleen O’Connor Duffany, Elizabeth C. Rhodes, Sara Rodonis, Andrea Aleaga, Gilma Galdamez, Andrea Tristán Urrutia, Rafael Pérez-Escamilla

## Abstract

Produce prescription programs (PRx) targeting different populations and conditions have been found to be effective. However, few have focused on pregnant women. The objectives of this study were to assess the impact of the Food4Moms (F4M) PRx on 1) healthy eating stages of change 2) intake of fresh produce, and 3) household food security among pregnant Latina women. F4M recruited low-income Latinas living in Greater Hartford, Connecticut that received a “produce prescription” from a Registered Dietitian based at the community-based organization (CBO) where the program was implemented. Participants were offered $100 per month for 10 months through Fresh Connect debit cards to purchase fresh produce from two food retailers or the equivalent value in fresh produce delivered at home. To be fully enrolled in F4M, participants had to complete a baseline survey and the first nutrition education interactive session. Enrolled participants were offered additional nutrition education sessions at the CBO and received text messages with nutrition tips as well as reminders to spend their remaining benefit balances towards the end of each month. A single-group pre-post study design was used to assess the impact of F4M 10 months after the card activation. No attrition bias was detected when comparing the characteristics of those completing (N=113) vs. those not completing the endline survey (N=41). Pre-post Wilcoxon signed-test or paired t-test analyses showed that F4M had a positive impact on healthy eating readiness (p < 0.001), the consumption of fruits (p < 0.001) and vegetables (p < 0.001), and household food security (p = 0.034). F4M is a promising community-engaged PRx program that may improve readiness for healthy eating, produce intake, and household food security. Implementation research is needed to find out how to effectively scale out and sustain programs like F4M. The study was registered in ClinicalTrials.gov (identifier: NCT05907616).

## Introduction

A higher intake of fruit and vegetables is consistently associated with a reduced risk of chronic diseases as well as lower all-cause mortality [1]. Despite these well-known benefits, intake levels in the United States remain suboptimal. According to the Center for Disease Control and Prevention, only 12.3% of adults meet the recommended fruit intake and 10% meet vegetable intake recommendations, with even lower consumption among individuals living in poverty (6.8%) [2]. Among pregnant women, fruit and vegetable intake is similarly inadequate. Data from the National Health and Nutrition Examination Survey (NHANES) indicate that only 35.4% meet recommended intake levels. On average, pregnant women consume 1.3 cups of fruits and 1.6 cups of vegetables per day [3], which falls short of the 2 to 2.5 cups of fruits and 2.5 to 3 cups of vegetables daily intake recommended by the Dietary Guidelines of Americans 2020-2025 [4]. Optimal nutrition before and during pregnancy is essential for maternal and fetal health. Adequate consumption of fruits and vegetables, rich in fiber, vitamin, minerals and antioxidants, has been associated with increased gut microbiota diversity, as well as a reduced risk of gestational diabetes, preeclampsia, and constipation problems [5] [6] [7]. Higher intake of fruit and vegetables has also been linked to improved self-management of gestational diabetes [8]. Maternal fruit and vegetable intake during pregnancy may further influence infant health outcomes as it may influence the infant gut microbiome at 2 months of age [9] and is positively associated with fetal and infant growth through six months [10]. At the same time, food security defined at the World Food Summit, as a condition in which “all people at all times have physical and economic access to sufficient, safe and nutritious food” [11], the lack of food security known as food insecurity remains a significant public health challenge, in 2024 13.7% of U.S. households were food insecure, with substantially higher prevalence among households below the poverty threshold (39.4%), single female headed households (36.8%), Black (24.4%), and Hispanic households (20.2%), and households with children (18.4%) [12], and between 10 and 14% of pregnant women in the US experience food insecurity [13] [14], which is associated with poorer diet quality and increased risk of adverse maternal and pregnancy outcomes [15] [16]. Furthermore, food insecurity is associated with limited access to nutritious and healthy foods including fruits and vegetables. More specifically, pregnant women experiencing food insecurity consume fewer vegetables [17]. A study conducted with low-income Hispanic pregnant women found that 43% reported low to very low food security and those with very low food security consumed a significantly lower variety of fruits and vegetables, likely because of reduced availability of fresh produce at home [18]. These findings are consistent with other studies linking food insecurity to poorer overall diet quality and fruit and vegetable intake among pregnant women [19]. Together, suboptimal fruit and vegetable intake and food insecurity during pregnancy represent critical, interrelated public health concerns with implications for both maternal and intergenerational health. Food assistance programs for pregnant women such as the Special Supplemental Nutrition Program for Women, Infants, and Children (WIC) improve perinatal outcomes, birth outcomes, access to nutritious foods, and prenatal care attendance [20]. Emerging evidence suggests that PRx programs are a complementary innovative strategy to improve access to healthy foods. PRx are interventions in which healthcare providers prescribe fruits and vegetables to patients, typically providing financial incentives or vouchers redeemable for fresh produce. PRx programs are likely to improve dietary behaviors, food security, and health outcomes. A systematic review of 20 PRx interventions conducted in the U.S. found positive impacts of PRx on food security and half of the studies also reported significant improvements in fruit and vegetable intake, along with improvements in blood glucose control, proxied by HbA1c, among individuals with type 2 diabetes [21]. PRx programs have also been found to increase fruit and vegetable consumption among low-income participants with poor cardiometabolic health when incentives are provided for at least six months [22]. Produce prescription programs that combine fruit and vegetable incentives with nutrition education have been shown to increase fruit and vegetable intake and food security [22]. Further evidence from the evaluation of the Vouchers 4 Veggies PRx program in San Francisco, California, demonstrated that monthly incentives of $20-$40 over a period of six months significantly improved fruit and vegetables consumption and food security in a diverse, low-income population [23]. A PRx program for pregnant African American women in Flint, Michigan, provided $15.00 fruit and vegetable prescription at each prenatal visit, redeemable at farmer markets and food hubs. The program significantly improved food security by late pregnancy and into the postpartum period, while fruits and vegetables intake increased during early to mid-pregnancy but declined after childbirth [24].

Produce prescription programs (PRx) targeting different populations and conditions have been found to be highly cost-effective [25]

While these findings underscore the potential of PRx programs to improve dietary intake, food security, and health outcomes, most studies have focused on individuals with chronic diseases and very few, have focused on low-income pregnant women despite of the promising nutritional and health benefits for this population. In addition, it is important to evaluate the influence of PRx programs on behavior change mediators, such as readiness to adopt and sustain healthy eating practices.

This paper is very timely since Congress recently passed the Consolidated Appropriations Act of 2026 through the Department of Health and Human Services, providing $15 million in grants for produce prescription for pregnant women [26]

### Objectives

The objectives of this study were to assess the impact of the Food4Moms (F4M) PRx on 1) healthy eating stages of change, 2) intake of fresh fruits and vegetables, and 3) household food security among pregnant Latina women.

## Materials and methods

This study was a collaboration between Wholesome Wave, the Hispanic Health Council and Yale Griffin Prevention Research Center and Yale School of Public Health.

Food4Moms was codesigned and tested for feasibility using community-engaged implementation science methods, the codesign phase included iterative refinements based on input through three consecutive listening sessions with 21 Latina women with low incomes who were either pregnant or had a child younger than two years and were residing in Greater Hartford, Connecticut. Following the codesign phase, the program was tested for feasibility with 20 participants. During this phase we identified implementation challenges and evaluated participant satisfaction levels. More detail on the codesign and feasibility testing of the program has been published elsewhere [27]. Findings from the feasibility phase led to the addition of more content on pregnancy related topics and infant feeding to the interactive nutrition education sessions. A text message was also added with the remaining benefit balance on the card close to the end of the month. Participants in the feasibility phase reported high satisfaction with all program components [27].

This paper describes the impact evaluation of F4M, program’s participants were recruited through a community-based organization, the Hispanic Health Council Maternal Health Programs, and the local Supplemental Nutrition Program for Women, Infants and Children (WIC) Office.

F4M’s impact was assessed following a single-group pre-post study design with 154 participants, which included those enrolled during feasibility phase. Throughout program design, implementation, and evaluation of F4M, community-engaged methods were applied through a person-centered lens to ensure cultural relevance, address structural barriers to healthy food access, and center the perspectives of participants.

### Conceptual frameworks

Two complementary frameworks guided program development, implementation, and evaluation across these three study phases. The Program Impact Pathway (PIP) framework was used to guide the codesign phase as well as the program implementation and evaluation phases [28]. The initial basic PIP framework for the F4M program outlined the main activities required during the study period to achieve the desired objectives and outcomes of improving produce intake and food security among study participants. These activities included: 1) community outreach; 2) screening and recruitment; 3) attendance to interactive nutrition education sessions provided by the Hispanic Health Council’s SNAP-Ed program; 4) prescription for and enrollment in the PRx; and 5) activation and redemption of incentives that were provided as a Fresh Connect debit card for the vast majority of participants. Each of these core PIP activities were further strengthened by the addition of supportive action identified during the codesign and feasibility phase of the study, including text messages and staff support with card activation and redemption challenges. These supportive actions were intended to address barriers and facilitate participant engagement, making it easier for participants to enroll in and actively participate in the program based on their input and experiences (Fig. 1)

**Fig. 1.**
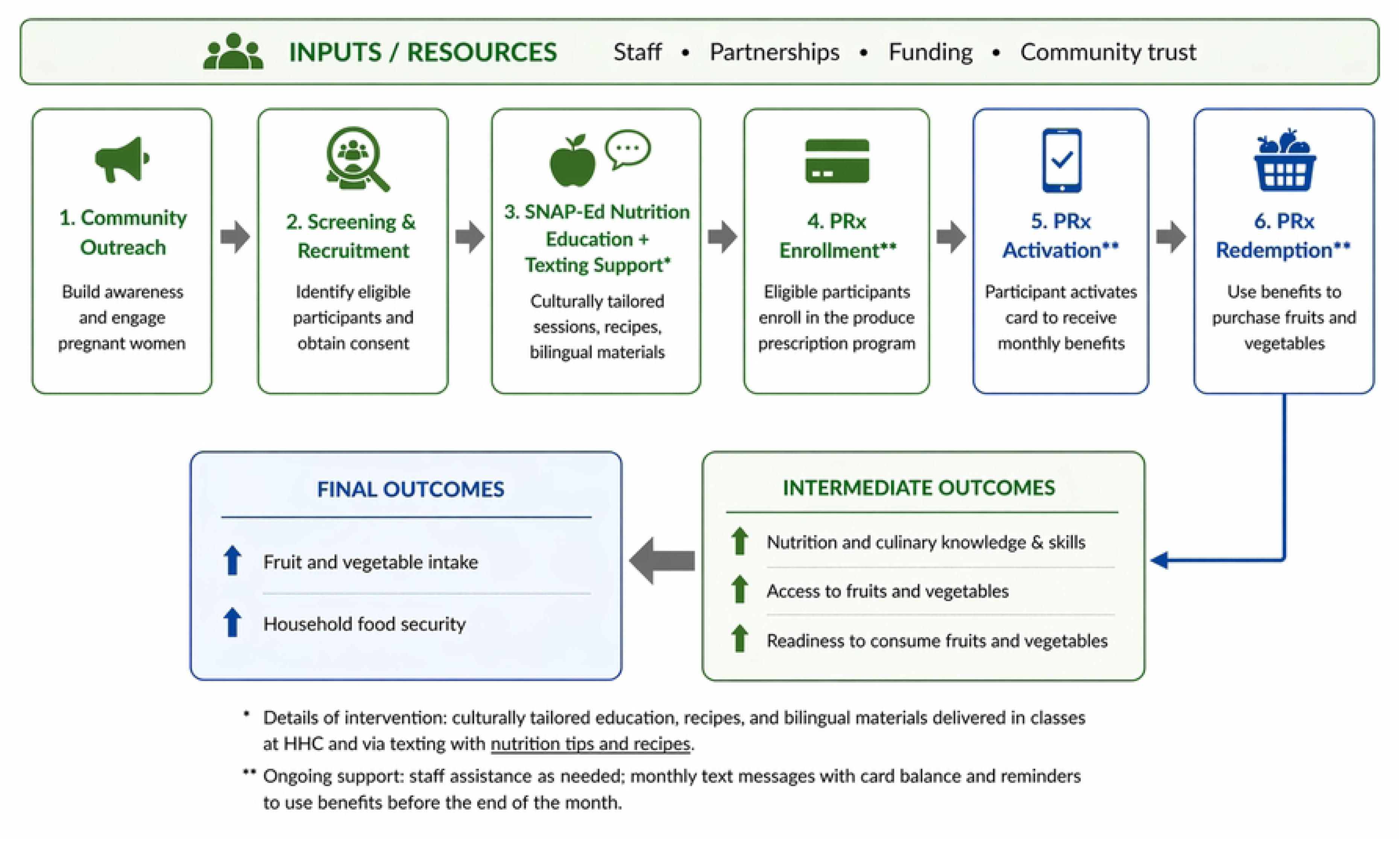
Program Impact Pathway (PIP) of the Food4Moms PRx implemented in Hartford, Connecticut.

The PIP illustrates the sequence of core program activities, from community outreach through incentive redemption, and their relationship to intermediate, and final outcomes, including improvements in fruit and vegetable readiness and intake, and in household food security. Supportive actions through staff and text messages identified during the codesign and feasibility phases were integrated across all stages to enhance participant engagement and reduce barriers to program participation.

The Capability, Opportunity, Motivation–Behavior (COM-B) model was used to further inform program design and confirm that the codesigned F4M program included the necessary components to support behavior change related to fruit and vegetable consumption (Table 1) [29]

**Table 1.**
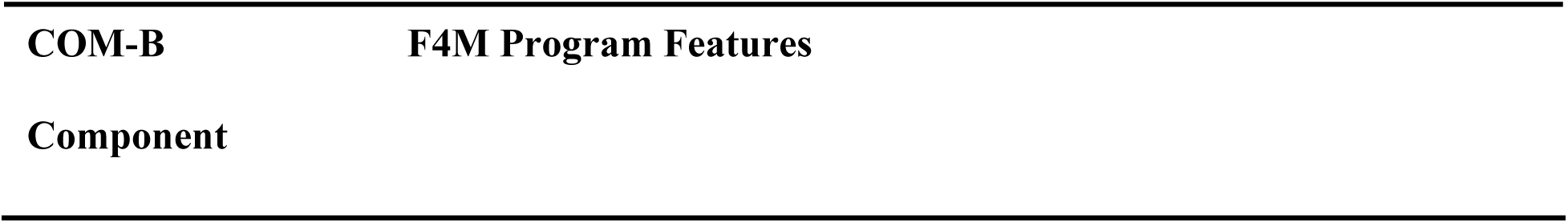
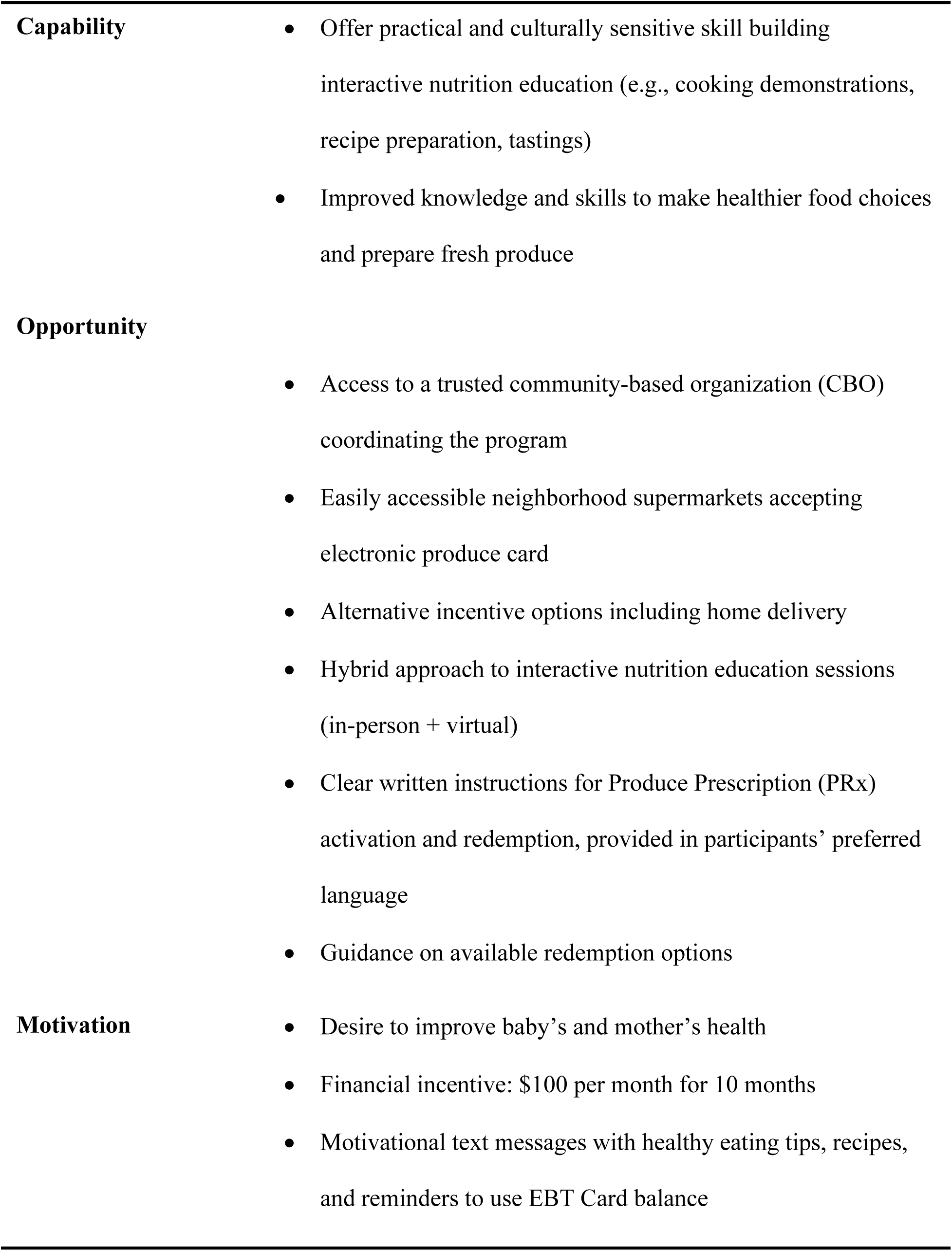
Mapping of Food4Moms produce prescription program components to the COM-B Framework.

According to the COM-B model, behavior change occurs when individuals have the capability, opportunity, and motivation to perform a given behavior [29]. Within the F4M program, capability was strengthened through the inclusion of practical and culturally sensitive interactive nutrition education sessions designed to improve participants’ knowledge and skills related to healthy eating and food preparation. These sessions aimed to increase participants’ ability to make healthier food choices and prepare fresh produce by incorporating vegetable recipe demonstrations and recipe tasting. Opportunity was supported through multiple program features that made participation easier and more accessible. Participants have access to a trusted community-based organization (CBO) coordinating the program, easily accessible neighborhood supermarkets accepting the electronic produce card, and an alternative option of home delivery. The hybrid approach to nutrition education sessions (in-person and virtual), clear verbal and written instructions for Produce Prescription (PRx) options, enrollment, activation and redemption provided in participant’s language of preference further facilitated engagement. Motivation was addressed by different factors since they were motivated by the desire to improve their baby’s and their own health, the financial incentive of $100.00 per month for 10 months to buy fresh produce, and motivational text messages providing healthy eating tips, recipes, and reminder to fully use their monetary incentives before the end of each month.

### Human subjects

This study was submitted as a single IRB to Yale University, indicating that the Hispanic Health Council and Wholesome Wave agreed that Yale University would provide the ethical and regulatory oversight for the study. IRB approval was received from Yale University on May 10, 2023 (IRB protocol ID: 2000034840), prior to study implementation. All partners, including the Hispanic Health Council and Wholesome Wave, helped prepare and review all materials submitted to the Yale University IRB and accepted its determinations.

### Study design

This study employed a single-group pre-post study design to assess the impact of the F4M produce prescription program on produce intake, household food security, and readiness for healthy eating Participants were assessed via surveys at baseline prior to program initiation and at endline which was within two months following completion of their ten-month participation in the study. A control group or comparison group was not included due to ethical considerations. Because the intervention provided financial incentives to purchase fresh produce to a highly food insecure population, withholding the intervention was deemed unethical since evidence demonstrates that financial incentives are effective in reducing food insecurity.

#### Eligibility criteria and recruitment

Participants were eligible for the study if they met the following criteria: identified as Latina, age 18 years or older, in their first or second trimester of pregnancy, resided in the Greater Hartford area in Connecticut, were English or Spanish speaking and were eligible for or currently participating in any one of the following programs WIC, SNAP, or Medicaid (state-based healthcare). Recruitment was conducted with bilingual (English/Spanish) flyers distributed by bilingual bicultural recruiters to local WIC offices, Hispanic Health Council Maternal and Child Health Programs, community outreach events and local community clinics offering prenatal, perinatal, postnatal and postpartum care services. After a brief explanation of the study, women who expressed interest were screened for eligibility either in person at the Hispanic Health Council or by phone using an online screening form to make sure they met the eligibility criteria.

#### Participant enrollment in the study

Eligible participants who agreed to participate provided verbal informed consent in their preferred language (English or Spanish) prior to baseline data collection. A research staff member called the participants to consent them over the phone, participants were provided access to the form electronically before or during the call so that they could follow along as the research staff explained the study. Once consented, all participants received a hard or digital copy of the consent form.

Consented participants were sent a link by text to complete a 20-minute baseline survey in their preferred language (English or Spanish) and invited to attend the first nutrition class in the format that best suited their needs, which included in person or virtual, English/Spanish language and various times (during work hours or after work hours). Participants were considered fully enrolled in the study after providing their consent, the baseline survey, and attending the first mandatory nutrition session; all participants were also invited to the remaining nutrition education sessions offered after completing enrollment (see Fig. 2).

**Fig. 2.**
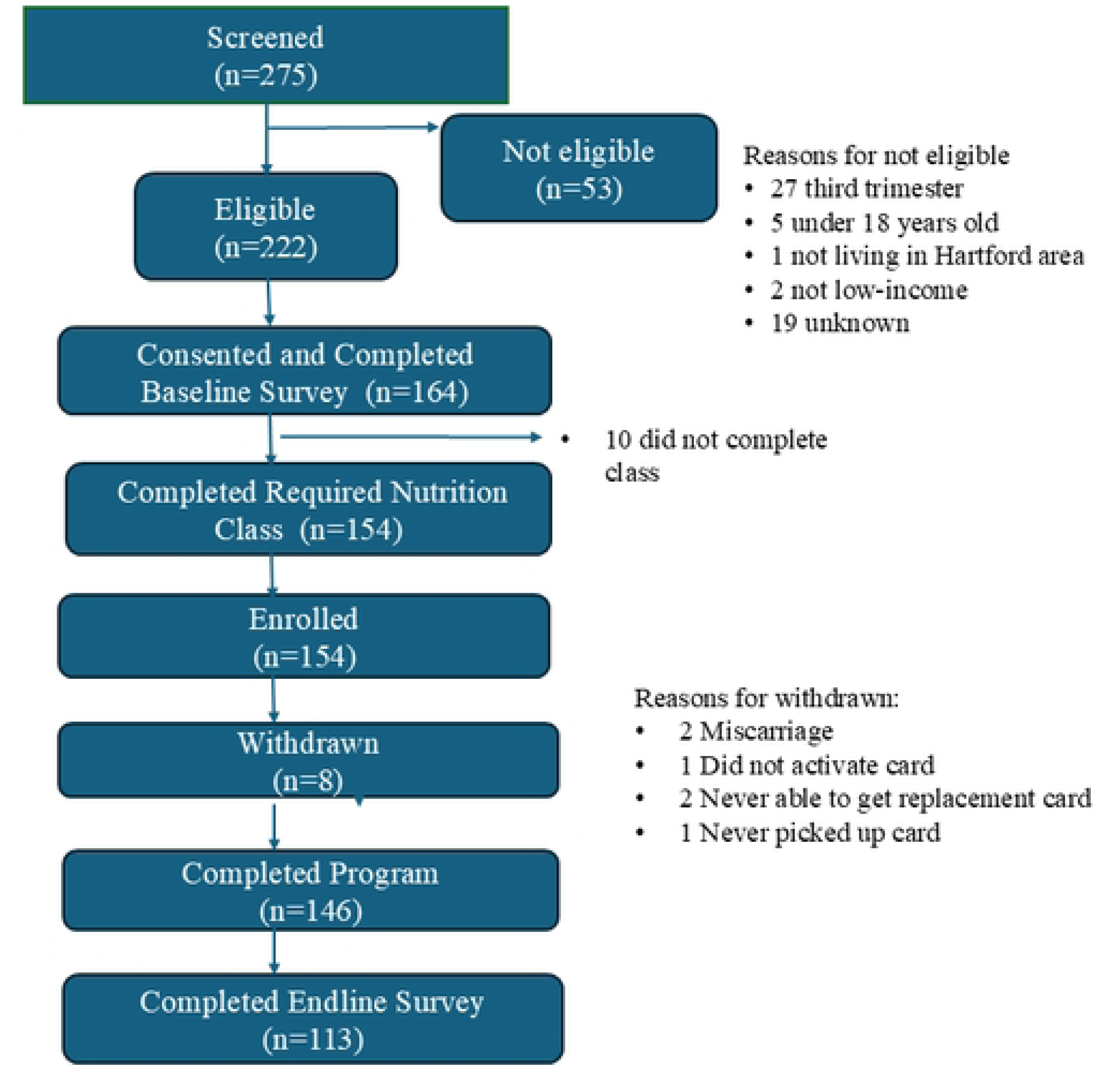
Flow chart of Food4Moms participants in program evaluation study in Greater Hartford, Connecticut.

At the conclusion of the first nutrition session, participants received a “produce prescription” signed by the F4M Registered Dietitian and selected their preferred method for receiving the monthly program benefits. Research staff explained the two options available for redeeming incentives, a Fresh Connect debit card or home delivery of fresh produce, both of which are described in the incentive section below.

The participant enrollment period occurred from July 24, 2023, through July 30, 2024. During this time, 275 individuals were screened, of whom 222 met the study’s eligibility criteria. A total of 164 eligible participants provided consent and completed the baseline survey (Fig. 2). Of these, 10 participants who had completed the baseline survey did not attend the mandatory nutrition class and therefore did not fulfill the enrollment criteria. All participants were sent a confirmation text of the remaining nutrition sessions that they chose, and a reminder text one day before each session took place. Consequently, 154 participants were fully enrolled in the study. During the study period, eight participants were withdrawn from the study including two due to miscarriage, and six due to card related issues such as one never activated card, two never activated replacement card, two were never able to get replacement card, and one did not pick up the card at the Hispanic Health Council (Fig. 2). Overall, 146 participants completed the 10-month study period, of whom 113 completed the post intervention survey. The impact analysis is based on the 113 participants that completed pre and post surveys (Fig. 2).

#### Incentives distribution options

The Fresh Connect Card was a restricted-use debit card that was earmarked for the purchase of fresh produce at two authorized retailers, Stop & Shop supermarkets and Walmart. Food4Moms provided monthly incentives of $100 for 10 months for a total of $1000. Incentives were preloaded onto the card at the beginning of each month. Unused funds did not roll over to subsequent months, meaning that any remaining benefit balance at the end of the month was forfeited and not added to the following month’s allocation.

Participants who selected the home delivery option received fresh produce through orders placed twice a month, one at the beginning and the other at the middle of the month using Instacart. Orders were submitted by the project director at Wholesome Wave, who selected a standard seasonal produce package with a value equivalent to the $100 monthly incentive, independent of delivery or service fees. This approach ensured that participants received an equivalent value of fresh produce regardless of the incentive redemption method selected. Participants receiving home delivery did not have the ability to choose the specific types of fresh produce included in the delivery but had the option to select an equal mix of fruits and vegetables or an order that had more vegetables or more fruit. Participants were told that they would be given the opportunity to change their selection at the 3-month mark of the study. Most participants selected the Fresh Connect debit card, but there were exceptions. We initially had three participants requesting the produce delivery box option; however, since we needed a minimum order of 20 boxes, we were unable to fulfill these selections. Those three participants were provided with the Fresh Connect card. Within a month we were able to develop a produce delivery option using Instacart. There were also some participants who had a very difficult time receiving the card in the mail resulting in ordering multiple replacement cards. As a result, two of those participants were provided with the Instacart produce delivery option while the Fresh Connect card issues were resolved. In January of 2025, the Fresh Connect Fidelity National Information Services (FIS) payment processor changed its banking partner and at the same time Fresh Connect moved from personalized cards to anonymous cards. Therefore, all participants still enrolled in the program after January 1^st^, 2025, needed new cards mailed to them. There were multiple issues with mailing new cards to participants resulting in participants going without cards for one to two months. In the interim period, we provided Instacart to the five participants affected so that they could have access to fresh fruits and vegetables.

#### Interactive nutrition education sessions

To be fully enrolled in the study, participants were required to complete the first nutrition education session in their preferred language (English/Spanish) and format of their preference (in-person or online). At the end of this mandatory session, participants were then enrolled to receive their “prescription” (i.e., incentives to purchase fruits and vegetables). After enrollment, participants were invited to the rest of the nutrition education sessions offered by the Hispanic Health Council SNAP-Ed program. The classes were delivered in person or virtually with morning and evening schedules, and in participant’s language of preference (English/Spanish). The interactive nutrition education curriculum was developed specifically for Food4Moms and was informed by the 2020 Dietary Guidelines for Americans, as well as community members feedback obtained during the codesign and feasibility phases. In addition, culturally sensitive sessions were integrated throughout the curriculum to reflect participants’ cultural food practices, beliefs, and preferences, including discussions addressing cultural perceptions of pregnancy and postpartum nutrition. All sessions included a vegetable-based recipe demonstration and lasted approximately 90 minutes. As previously mentioned, after completion of the initial feasibility phase, an additional fifth session focused on infant feeding recommendations during the first year of life was incorporated into the curriculum. The nutrition education sessions focused on Healthy Eating during Pregnancy (mandatory); Dietary strategies for the prevention and management of pregnancy complications; Making Healthy Food Choices Using the Nutrition Facts Label; Food Safety and Practical Nutrition Tips; and Maternal Nutrition and Infant Feeding during the first year of lif**e** (Table 2).

**Table 2.**
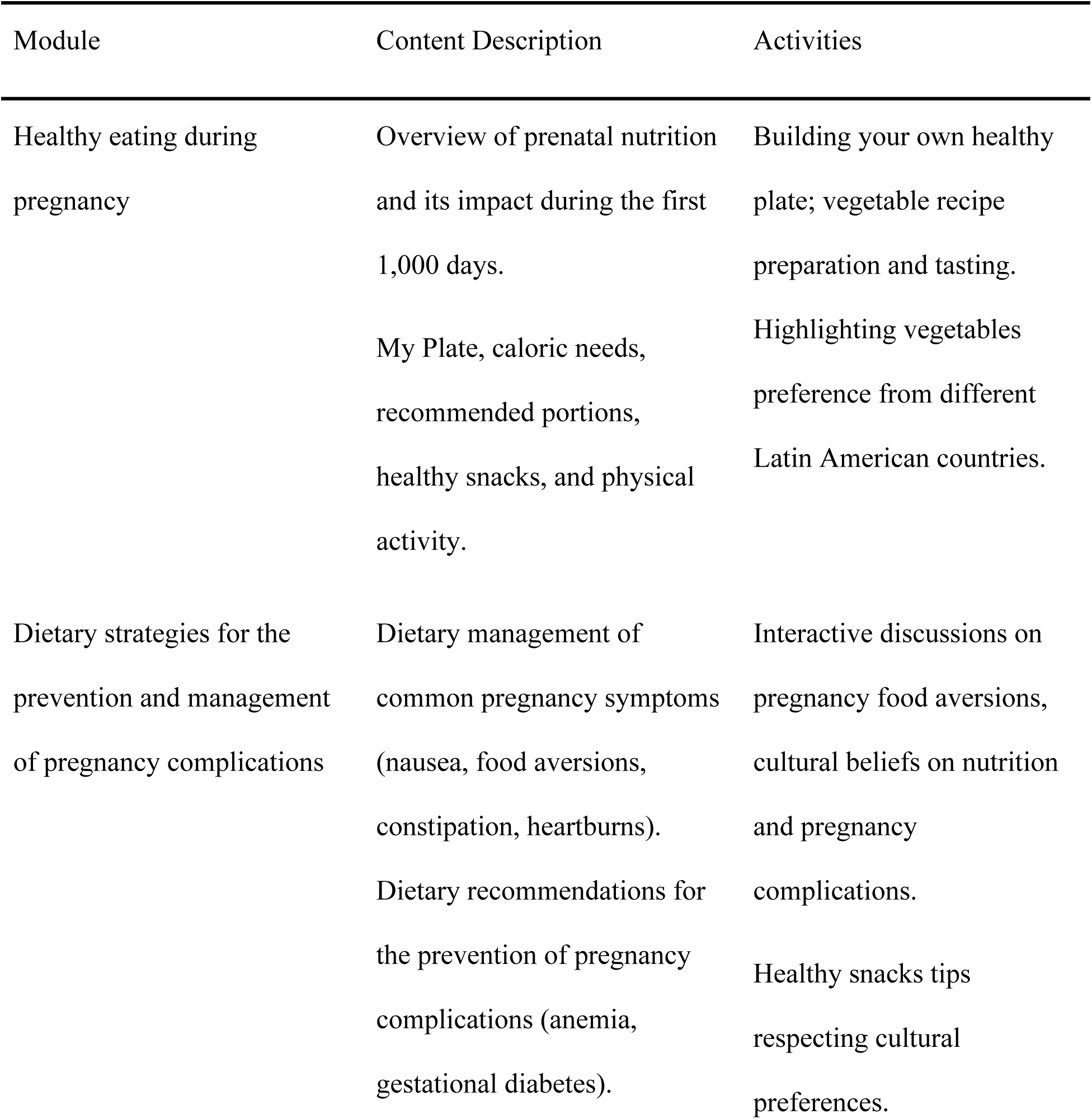
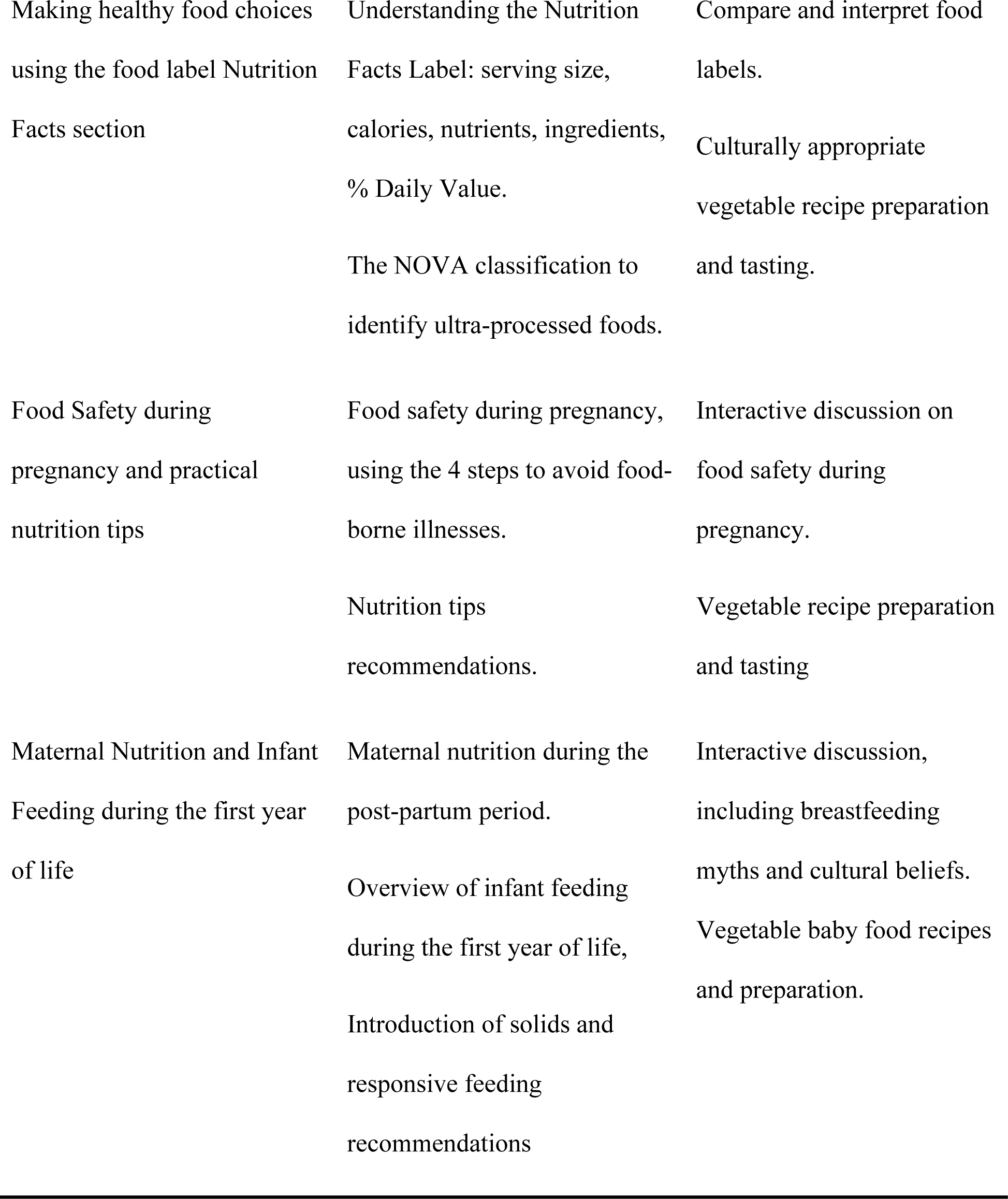
Content of Food4Moms bilingual and interactive nutrition education sessions delivered by community nutritionists to pregnant Latinas in Hartford, Connecticut.

Throughout the program, all participants received weekly text messages with nutrition tips, links to recipes, plus monthly reminders to use their produce incentives.

### Data collection

#### Survey instruments

A baseline survey and an endline survey were administered through an electronic link sent to participants via text. The surveys included pre-validated scales in English and Spanish, and those pertaining to this paper are described in the data collection section. Participant’s sociodemographic data at baseline (n=154) and among those who completed both pre and posttest (n=113) included: participant’s age, spoken language, household size, educational attainment, monthly income, employment status, number of children in household, country of origin, ethnicity, race. We collected data on participation during the last 12 months in the following government food assistance and health coverage programs: (1) the Special Supplemental Program for Women, Infants, and Children (WIC), (2) the Supplemental Nutrition Assistance Program (SNAP); and (3) Medicaid.

#### Baseline survey

The baseline survey took approximately 30-45 minutes to complete. Participants could complete the survey independently online (77%), have it administered by a trained bilingual and bicultural interviewer via phone or in person (9.1%) or in person but reading the questions to themselves (9.7%). The survey collected socio-demographic information; barriers to fruits and vegetables consumption; maternal physical health and pregnancy-related complications; main sources of nutrition knowledge during pregnancy; planned infant feeding practice; state of change for healthy eating, self-efficacy related to fruit and vegetable consumption and eating habits; a food frequency questionnaire focusing on fruit and vegetable consumption; household food security and participation on food assistance programs.

#### Endline survey

The endline survey had similar duration as the baseline survey, and included all measures from the baseline survey, along with additional postpartum questions related to maternal and infant health, pregnancy outcomes, infant feeding practices, and participant satisfaction with the program. The survey was intended to be completed within 2 months of program completion. Approximately 80% of the participants completed the endline survey within this timeframe, while the remaining 20% took longer to complete it.

### Data variables

#### Produce intake

Fruit and vegetable intake were assessed at baseline and endline using 10 items from the Dietary Screener Questionnaire assessment tool developed by the National Cancer Institute [30]. This validated, self-administered tool asks participants how often they consumed fruits and vegetables over the past month, with responses converted to cups/day.

#### Stage of change for healthy eating behaviors

Stages of changes or readiness for healthy eating were assessed with a standardized questionnaire based on the Transtheorical Model, which classifies individuals into the precontemplation, contemplation, preparation, action, or maintenance stages [31]

#### Household food insecurity

Food security was assessed at baseline and endline using the six-item short form of the U.S. Household Food Security Survey Module (USDA ERS). An additive score was computed based on the number of items affirmed. Participants’ scores were classified into one of the following mutually exclusive four categories using the recommended cut-off points: food secure (score –0); marginal food security (score =1), 2 to 4 – low food security (score 2-4) and very low food security (scores 5-6) [32]

### Statistical analyses

All data were analyzed using IBM SPSS for Windows® (V. 21) [33]. We generated socio-economic and descriptive characteristics at baseline for all the samples and compared them (N=154) with endline surveys. We conducted an attrition bias analysis by comparing the characteristics of those completing (N=113) vs. those not completing the endline survey (N=41), using the Chi-Square test. We compared the stages of change for healthy eating responses and the household food insecurity classification at baseline and endline using the non-parametric Wilcoxon sign-rank. We compared the proportion affirming at least one item of the Household Food Security scale at both time points with the Wilcoxon test. The cup equivalents of vegetable and fruits intake were compared at baseline and endline using the paired t-test. Statistical significance was set at a p-value <=0.05. Marginal significance was considered at a p value >0.05-0.10. Multivariable analyses were not necessary because there was no evidence of attrition bias among participants that completed the endline survey compared with those that didn’t.

## Results

### Sample characteristics

As shown in Table 3, 64.9% of the 154 enrolled participants chose to complete the baseline survey in Spanish, and a similar proportion (59.7%) were between 5 and 20 weeks of pregnancy.

**Table 3.** Baseline descriptive characteristics of Food4Moms participants, including comparisons with those that completed the endline survey.

|  |  | Baseline-All |  | Baseline-<br>endline<br>survey<br>completers |  | P value<sup>1</sup> |
| --- | --- | --- | --- | --- | --- | --- |
|  |  | N | % | N | % |  |
| Language answering<br>survey | English | 54 | 35.1% | 40 | 35.4% | .886 |
|  | Spanish | 100 | 64.9% | 73 | 64.6% |  |
| Weeks of pregnancy | 5-12 weeks | 35 | 22.7% | 25 | 22.1% | .764 |
|  | 13-16 weeks | 32 | 20.8% | 26 | 23.0% |  |
|  | 17-20 weeks | 25 | 16.2% | 18 | 15.9% |  |
|  | 21-24 weeks | 26 | 16.9% | 17 | 15.0% |  |
|  | 25-32 weeks | 16 | 10.4% | 11 | 9.7% |  |
|  | Don't Know | 20 | 13.0% | 16 | 14.2% |  |
| Age group | 18 -25 | 55 | 35.7% | 37 | 32.7% | .492 |
|  | 26 - 30 | 46 | 29.9% | 36 | 31.9% |  |
|  | 31-40 | 48 | 31.2% | 37 | 32.7% |  |
|  | 41 or higher | 5 | 3.2% | 3 | 2.7% |  |
| Respondent's Birth | Puerto Rico | 31 | 22.5% | 13 | 16.5% | .054 |
| Country/Territory | Peru | 26 | 18.8% | 21 | 20.4% |  |
|  | United States | 17 | 12.3% | 13 | 12.6% |  |
|  | Dominican Republic | 15 | 10.9% | 10 | 9.7% |  |
|  | Guatemala | 10 | 7.2% | 8 | 7.8% |  |
|  | Other (specify)<sup>2</sup> | 39 | 28.3% | 34 | 33.0% |  |
|  | Less than HS | 21 | 14.9% | 16 | 15.1% | .160 |
|  | diploma or equivalent |  |  |  |  |  |
|  | High school/GED | 110 | 78.0% | 85 | 80.2% |  |
|  | completed or some |  |  |  |  |  |
|  | college |  |  |  |  |  |
|  | BS degree or higher | 10 | 7.1% | 5 | 4.7% |  |
| Monthly income | Less than \$1,000 | 63 | 42.6% | 46 | 42.2% | .797 |
| | \$1,000 to \$1,999 | 49 | 33.1% | 35 | 32.1% | |
| | More than \$2,000 | 36 | 24.3% | 28 | 25.7% | |
| Employment status | Working full time or | 53 | 34.6% | 36 | 32.1% | .713 |
|  | part time |  |  |  |  |  |
|  | Unemployed/Student/<br>Home maker | 86 | 56.2% | 66 | 58.9% |  |
|  | Disability/SSI/SSDI<sup>3</sup> | 3 | 2.0% | 2 | 1.8% |  |
|  | Other | 11 | 7.2% | 8 | 7.1% |  |
| Access to a reliable<br>car | Yes | 89 | 57.8% | 64 | 56.6% | .873 |
|  | No | 38 | 24.7% | 29 | 25.7% |  |
|  | Sometimes | 27 | 17.5% | 20 | 17.7% |  |
| Household Food<br>Security Category | High food security | 29 | 21.6% | 22 | 22.7% | .882 |
|  | Marginal food<br>security | 18 | 13.4% | 14 | 14.4% |  |
|  | Low food security | 67 | 50.0% | 47 | 48.5% |  |
|  | Very low food<br>security | 20 | 14.9% | 14 | 14.4% |  |
| Enrolled in WIC in<br>previous 30 days | No | 51 | 33.1% | 38 | 33.5% | .823 |
|  | Yes | 103 | 66.9% | 75 | 72.8% |  |
|  | No | 104 | 67.5% | 81 | 71.7% | .152 |
|  | Yes | 46 | 29.9% | 30 | 26.5% |  |
| Household received SNAP benefits in past 30 days | Don't know/Prefer not to answer | 4 | 2.6% | 2 | 1.8% |  |
| Perceived general health status | Poor or Fair | 48 | 31.4% | 35 | 31.3% | .986 |
|  | Good | 70 | 45.8% | 51 | 45.5% |  |
|  | Very Good or Excellent | 35 | 22.9% | 26 | 23.2% |  |

<sup>1</sup> When comparing those with vs. those without completed endline surveys with Chi-square test, none of the differences were statistically significant or marginally significant.
<sup>2</sup> Less than 10 participants were born in the following countries: El Salvador (n=8), Mexico (n=8), Venezuela (n=5), Colombia (n=4), Nicaragua (n=2), Brazil (n=1), Ecuador (n=1), Italy (n=1), Panama (n=1), There were 2 participants who misunderstood the question thinking that it referred to the place of birth of the baby.
<sup>3</sup> SSI: Supplemental Security Income; SSDI: Social Security Disability Insurance

Most participants (78%) had completed high school, 87.7% were born outside of the continental United States, and only 34.6% reported being employed. Additionally, 75.7% reported a monthly income at or below $1999, and only 57.8% always had access to a car. Only 21.6% of participants experienced high food security, while 13.4% experienced marginal food security, 50% low food security, and 14.9% very low food security. Additionally, 29.9% of the participants were receiving SNAP benefits, 66.9% were enrolled in WIC, and 68.7% reported their health as good to excellent at baseline. There were no significant differences in socioeconomic characteristics between participants who completed the endline surveys and those who did not.

### Healthy Eating Readiness

Table 4 shows that the change between baseline and endline shifted toward more advanced stages of change for healthy eating was statistically significant (Wilcoxon signed-rank test, p < .001).

**Table 4.** Stages of change for current eating habits stages of change among Food4Moms participants in Hartford, Connecticut^1 N=113^.

|  | Baseline |  | Endline |  |
| --- | --- | --- | --- | --- |
|  | N | % | N | % |
| I have not been eating healthy, and I do not intend on changing my eating habits in the near future. | 3 | 2.7 | 3 | 2.7 |
| I intend to eat healthier in the next six months. | 51 | 45.1 | 20 | 17.7 |
| I intend to eat healthier in the next month. | 20 | 17.7 | 20 | 17.7 |
| I have been eating healthier in the last six months. | 32 | 28.3 | 38 | 33.6 |
| I have been eating healthy for more than six months. | 7 | 6.2 | 32 | 28.3 |

<sup>1</sup>Wilcoxon Signed ranks test, $p < 0.001$

At baseline, most participants were in earlier stages with 45.1% reporting intention to eat healthier within the next six months and 17.7% within the next month. A lower percentage of participants were in the action stage (28.3%) or maintenance stage (6.2%) for eating healthy. Pre-post analyses show that the proportion of participants that were at the action stage for healthy eating increased from 28.3% to 33.6%, and a larger increase was observed in the maintenance stage rising from 6.2% to 28.3%.

### Produce intake

Among 113 participants with both a baseline and endline survey, 101 had completed fruit and vegetable intake data. Food4Moms was positively associated with an increase in fruit and vegetable intake (Table 5).

**Table 5.**
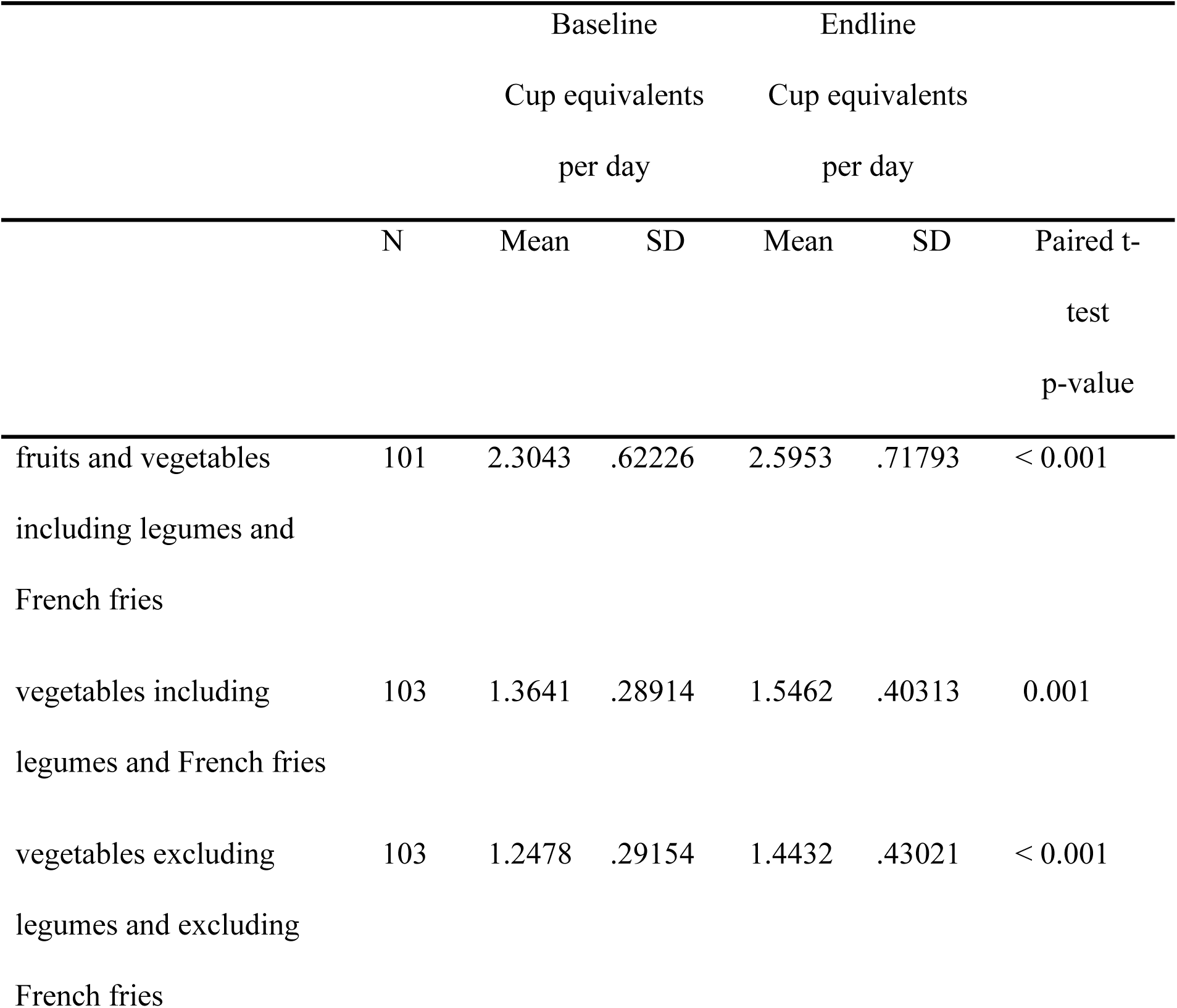
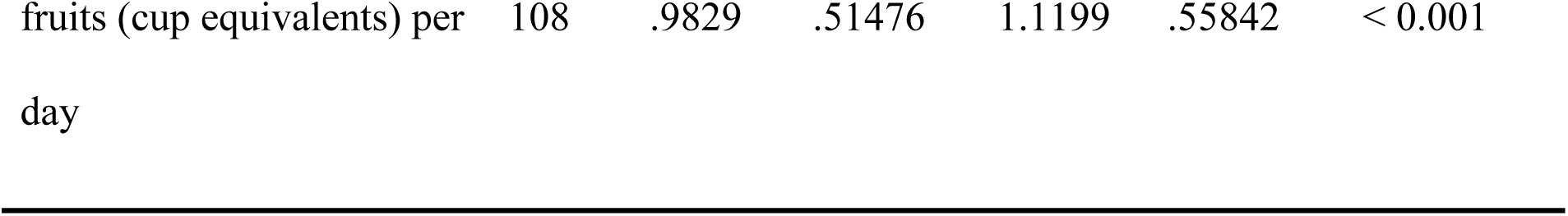
Changes in produce intake among Food4Moms participants in Hartford, Connecticut.

Total fruit and vegetable consumption (including legumes and French fries) increased from a mean of 2.30 to 2.59 cup equivalents per day (p < 0.001). Consumption of vegetables (including legumes and French fries) increased from 1.36 to 1.54 cup equivalents per day (p=0.001), while vegetable intake excluding legumes and French fries increased from 1.24 to 1.44 cup equivalents per day (p < 0.001). Likewise, fruit intake increased from 0.98 to 1.11 cup equivalents per day (p < 0.001).

### Household food insecurity

Table 6 presents changes in household food insecurity among Food4Moms participants from baseline to endline.

**Table 6.**
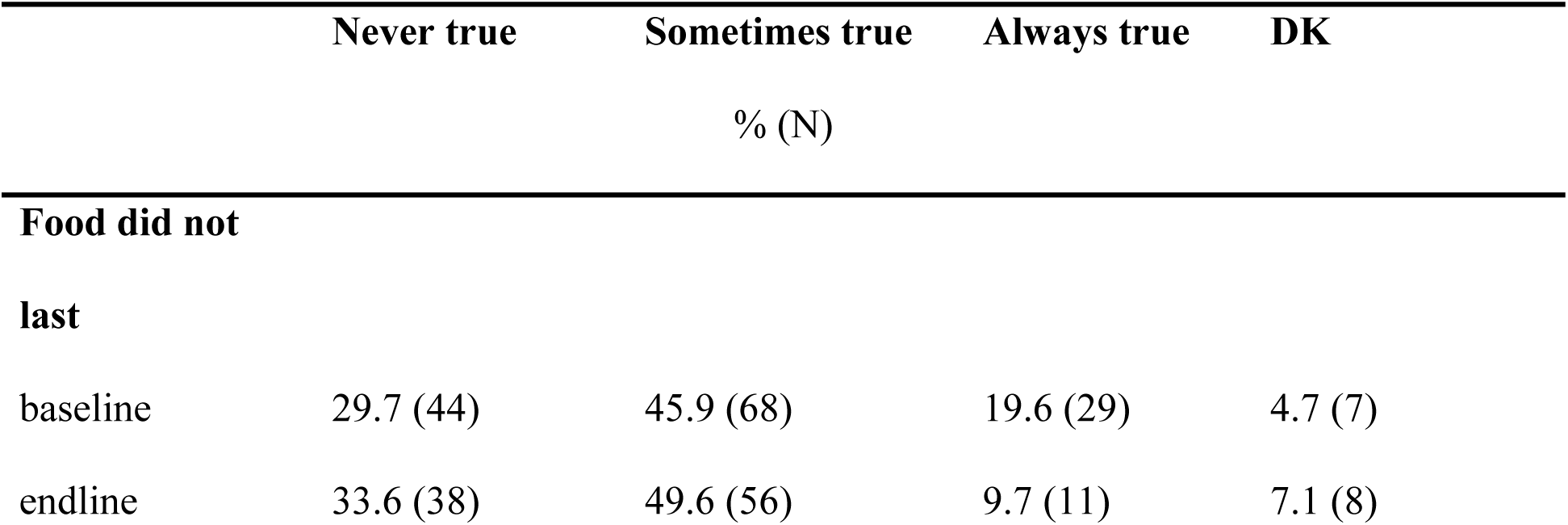
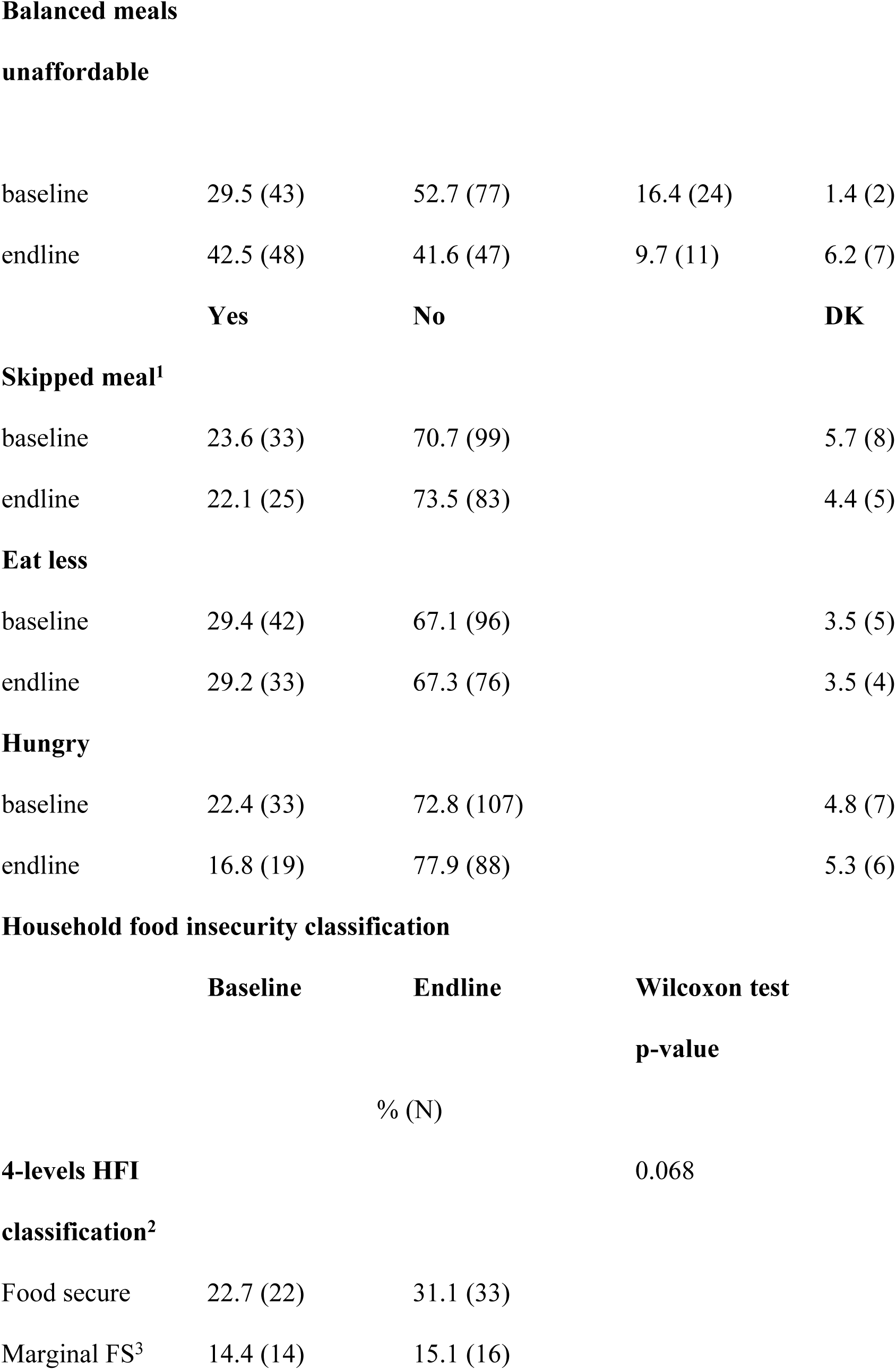
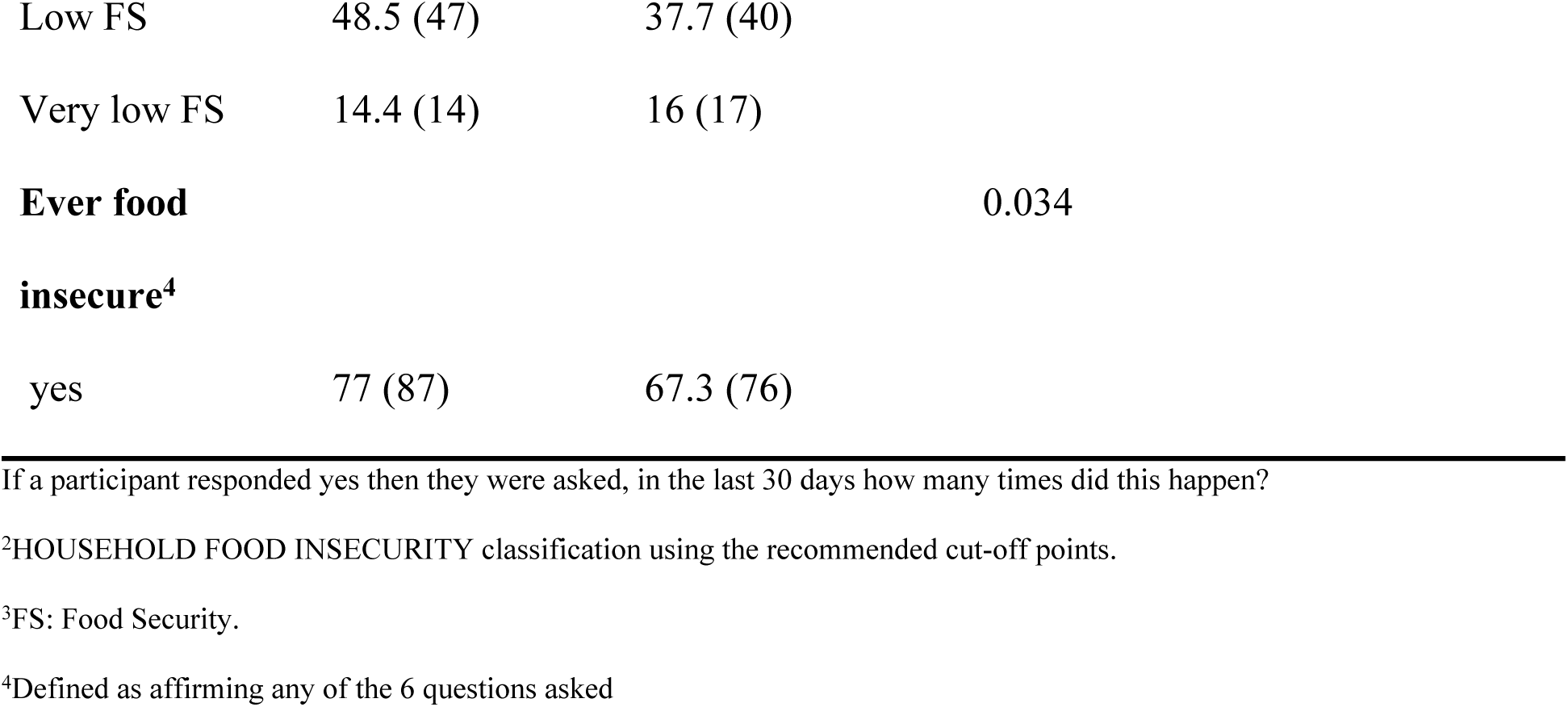
Household food insecurity among Food4Moms participants in Hartford, Connecticut. Assessed with the 6-item US Household Food Security Survey Module.

The proportion of participants reporting that food did not last as “always true” decreased from 19.6% at baseline to 9.7% at endline, while those reporting this as “never true” increased from 29.7% to 33.6%. Similarly, the proportion of participants reporting that balanced meals were unaffordable as “always true” decreased from 16.4% to 9.7% with a corresponding increase in those reporting “never true” from 29.5% to 42.5%. Changes in more severe food insecurity behaviors were modest. The proportion of participants reporting skipping meals decreased slightly from 23.6% to 22.1%, and those reporting being hungry but not eating decreased from 22.4% to 16.8%), while reports of eating less remained largely unchanged.

Overall, household food insecurity improved modestly. The proportion of participants classified as food security increased from 22.7% to 31.1%, while those with low food security decreased from 48.5% to 37.7%, and this association was marginally significant (Wilcoxon test, p = 0.068). Furthermore, the proportion of participants experiencing any food insecurity as proxied by affirming at least one of the six scale items decreased significantly from 77.0% to 67.3% (Wilcoxon test, p = 0.034).

## Discussion

Most PRx programs are delivered through healthcare systems, in contrast, F4M represents a person-centered, community engaged model designed specifically for low-income pregnant women coordinated by a community-based agency. Its quality assurance program enabled timely identification and resolution of implementation challenges with input from all partners, including program participants. In F4M the “prescription” was provided by the agency’s Registered Dietitian as part of a SNAP-Ed Program run by the Hispanic Health Council. This approach likely helped reduce barriers to participation, such as limited access to healthcare, transportation challenges, distrust of clinical systems, while leveraging trusted community relationships to improve program reach and engagement. This is especially true when serving hard to reach population, particularly recent immigrants that may not qualify for food assistance programs; notably, in this study most participants were born outside of continental United States.

Participation in F4M was positively associated with improved readiness for healthy eating, and a higher intake of both vegetables and fruits. This suggests that changes in readiness to adopt healthy eating behaviors translate into measurable dietary improvements.

The findings from our study align with prior PRx research with other target groups, demonstrating improvements in diet quality and food security. For example, a pooled multisite evaluation of nine Wholesome Wave PRx programs across 12 U.S. states (n=3,881), tree focused on 2-18 years old children and six on adults with cardiometabolic diseases found significant increases in fruit and vegetable intake, as well as improvements in food security and self-reported health among low-income participants receiving produce incentives through vouchers or electronic cards [34]. Similarly, a Canadian healthcare-based PRx program providing weekly vouchers alongside nutrition education over 12 months to adult women and men reported reductions in food insecurity, increased fruit and vegetable intake, and improvements in select clinical biomarkers, including triglycerides, fasting insulin, and vitamin C levels [35].

Additional evidence from pediatric and family-based Prx program further supports these findings. A scoping review of 19 pediatric PRx studies providing vouchers or food boxes found a reduction of food insecurity in five out of the six studies assessing changes in food insecurity and increases in fruit in vegetables intake in five out of the seven studies measuring dietary intake [36]. Qualitative and mixed-methods studies also reported broader benefits, including reduced financial strain, improved nutrition knowledge, and enhanced cooking practices, particularly when programs included home delivery or were integrated into existing community services [37,38].

Despite this growing body of evidence, there is a dearth of evidence on PRx programs specifically targeting pregnant women. This represents a critical gap addressed by our study, given the heightened nutritional needs during pregnancy and the potential for intergenerational health impacts. One notable example is a PRx program implemented among pregnant African American women in Flint, Michigan, which provided $15 fruit and vegetable prescriptions at prenatal visits. That program improved food security through pregnancy and into the postpartum period; however, increases in fruit and vegetable intake observed during pregnancy were not sustained after childbirth [24]. These findings underscore both the promise of PRx interventions during pregnancy and the need for continued support in the postpartum period.

F4M builds on and extends this limited evidence base by targeting a predominantly Hispanic, immigrant population and integrating culturally tailored interactive nutrition education, recipe demonstrations, and ongoing engagement through text messaging. To our knowledge, few PRx programs for pregnant women have combined financial incentives with comprehensive, culturally responsive nutrition education delivered in a community setting. This integrated approach may be particularly important for sustaining behavior change beyond pregnancy. Our findings are consistent with a recent systematic review of Food is Medicine programs for pregnant women, which found improvements in diet quality, food security, and some birth outcomes, our findings support the potential of PRx interventions during this critical life stage. However, the review also highlighted substantial heterogeneity in program design and outcomes, as well as a lack of standardized measures and long-term follow-up, limiting conclusions about overall effectiveness [22]

Taken together, our findings contribute to a small but growing literature suggesting that PRx programs for pregnant women can improve dietary behaviors and food security. They also highlight the potential added value of community-based, culturally tailored models in reaching underserved populations and addressing structural barriers to healthy eating.

An important limitation of this study is the use of a one-group pre–post design without a comparison group, which limits our ability to establish causality and disentangle program effects from external influences. However, the observed improvements in readiness for healthy eating—a plausible mediator of the relationship between F4M participation and increased fruit and vegetable intake—provide supportive evidence that the intervention contributed to the dietary changes observed. The absence of a control group was intentional and guided by ethical considerations raised by community partners, who did not feel it was appropriate to withhold access to fresh produce from low-income pregnant women. In addition, available funding constrained the study design to a single-group evaluation. Given that the primary objectives were to codesign the intervention, assess implementation feasibility, and estimate its potential impact, this approach was appropriate for this phase of research [27], which is consistent with other FIM PRx studies [24]

Building on these findings, we recommend future research based on a randomized controlled trial design to evaluate the effectiveness of F4M more rigorously. We specifically recommend comparing different levels of monthly financial incentives rather than including a no-intervention control group, thereby maintaining ethical integrity while generating evidence to inform scalable program design. Identifying an optimal and cost-effective incentive level will be critical for broader policy translation and program implementation and dissemination.

This study also has several notable strengths. First, it was grounded in a community-engaged, codesign approach, ensuring that the intervention was responsive to participants’ needs and preferences. Program implementation was guided by a quality assurance framework informed by the Program Impact Pathways (PIP) framework and the COM-B model, alongside iterative feedback collected through listening sessions and surveys with participants. This allowed for continuous refinement and adaptation of the program.

Second, the study was implemented by the Hispanic Health Council, a trusted community-based organization with more than four decades of experience serving Hispanic populations. Its longstanding presence and established relationships within the community were instrumental in engaging participants, particularly recent immigrants who are often underrepresented in research. The organization’s history of community-based participatory research further strengthened the relevance and acceptability of both the intervention and the evaluation methods.

Third, F4M leveraged an existing, well-established SNAP-Ed program to deliver its interactive nutrition education component. This integration ensured that the intervention was not only evidence-based but also culturally and linguistically appropriate, as well as person-centered in its delivery. As a result, both the program and the research processes were aligned with the cultural context and lived experiences of participants.

Fourth, the partnership with Wholesome Wave provided critical infrastructure for the activation and tracking of Fresh Connect monthly benefits, supporting accurate monitoring of program utilization. Finally, collaboration with an academic partner, the Yale School of Public Health, ensured methodological rigor in study design, data collection, and analysis, strengthening the validity and credibility of the findings.

Moving forward, implementation research is needed to determine how programs like F4M can be effectively scaled and sustained at state and national levels. There is a need to identify the level of financial incentives required to meaningfully improve fruit and vegetable intake while remaining cost-effective and feasible for government agencies and other funders. The optimal duration of benefit provision also warrants further investigation, especially given that infants are recommended to begin complementary feeding around six months of age and could directly benefit from increased household access to healthy foods.

In addition, future studies should evaluate this model across diverse racial and ethnic populations and in larger-scale effectiveness prospective studies with longer follow-up periods. Such research is essential to assess not only dietary outcomes, but also the impacts of programs like F4M on pregnancy, birth, infant feeding, and child health and development outcomes.

Generating this evidence will be critical to inform policy decisions and support the broader integration of produce prescription programs into comprehensive maternal and child health strategies.

## Conclusion

Food4Moms is a promising person- and family-centered prescription community-engaged program that may improve readiness for healthy eating, produce intake, and household food security.

## Data Availability

While the manuscripts of this study are being published the data set and codes will be made available by the senior author (RP-E) upon reasonable request.

## Acknowledgements

The authors deeply thank all the staff from the SNAP-ED program for their contributions to the interactive nutrition education sessions. They are also very grateful to all the mothers, babies, and their families who participated in the program.

